# Risk factors for ICU admission due to severe acute respiratory infection in Brazil: A *multicenter case-control study using national data*

**DOI:** 10.1101/2025.11.20.25340681

**Authors:** Ryan R.O. de Paula, Priscila R. Cintra, Eric Francelino Andrade, Luiz O.O. Pala, Ramiro Mendonça Murata, Luciano J. Pereira

## Abstract

**Objective:** To identify risk factors associated with Intensive Care Unit admission due to Severe Acute Respiratory Infection in Brazil between 2021 and 2023, across different geographical levels during the COVID-19 pandemic.

**Methods:** A retrospective case-control study was conducted using data from the OpenDataSUS system. Logistic regression models, within the framework of Generalized Additive Models for Location, Scale, and Shape, incorporated demographic, clinical, and vaccination variables. Model performance was assessed through cross-validation.

**Results:** Male sex, age between 60 and 70 years, dyspnea, respiratory distress, oxygen saturation <95%, diabetes, and obesity significantly increased the risk of Intensive Care Unit admission admission. Cough and sore throat were protective factors in the national, regional, and state-level analyses. COVID-19 and Influenza vaccination were consistently associated with reduced risk across all geographical levels, particularly among elderly patients. At the municipal level (Lavras, Minas Gerais), obesity had the greatest impact, while fever and odynophagia emerged as additional predictors, suggesting the influence of local factors such as socioeconomic conditions and hospital infrastructure. The models demonstrated moderate performance (AUC: 0.60–0.81).

**Conclusions:** Severe respiratory symptoms, comorbidities, and male sex are key determinants of Severe Acute Respiratory Infection -related Intensive Care Unit admission. COVID-19 and Influenza vaccination showed a protective effect, especially in older adults, which may explain the observed reduction in risk after age 60. Regional differences underscore the importance of considering sociodemographic and structural factors in health planning and resource allocation.

## Introduction

Severe Acute Respiratory Infection (SARI) is characterized by signs and symptoms caused by infectious agents such as influenza, respiratory syncytial virus (RSV), parainfluenza, and adenovirus [1]. Patients typically present with influenza-like symptoms (cough, sore throat, fever, rhinorrhea, nasal congestion), often accompanied by myalgia, headache, or arthralgia. These may be associated with severity indicators such as dyspnea, tachypnea (≥20 breaths/min), hypoxemia, oxygen saturation <95% on room air, clinical worsening of an underlying condition, hypotension, or acute respiratory failure, occurring during seasonal periods and in the absence of another specific diagnosis [2].

Due to its epidemiological relevance, SARI must be reported and investigated by the epidemiological surveillance system through the Notifiable Diseases Information System (SINAN). This enables the prevention and monitoring of severe cases, the identification of circulating viruses, antigenic analyses, the monitoring of new variants, and the control of outbreaks, thereby supporting public health decision-making [2].

This surveillance became even more essential during the COVID-19 pandemic, caused by Severe Acute Respiratory Syndrome Coronavirus 2 (SARS-CoV-2), one of the greatest public health challenges of the century [3]. As of September 19, 2024, Brazil had confirmed 38,915,370 cases and 713,205 deaths, resulting in a case fatality rate of 1.8% and a mortality rate of 339.4 per 100,000 inhabitants [4]. Risk factors for greater severity include advanced age, comorbidities (obesity, diabetes, cardiovascular diseases), male sex, and lack of vaccination [1,5]. The association with chronic diseases is linked to persistent inflammation, with elevated levels of pro-inflammatory cytokines and an imbalance of the renin–angiotensin system, which increases ACE2 expression and favors a pro-inflammatory state [6].

Demographic and regional factors also influence SARI outcomes. In Pernambuco, population vulnerability and a low Municipal Human Development Index (MHDI) compromised healthcare services and hindered the transfer of severely ill patients [3]. In Pará, the limited availability of ICU beds may have explained variations in admissions, highlighting the interaction between socioeconomic factors and pathophysiological processes [6].

Retrospective observational studies, such as case-control studies, allow the evaluation of multiple risk factors for a given outcome. They are useful for analyzing public health interventions such as vaccination campaigns, treatment protocols, and control policies, as well as for comparing patients exposed or not exposed to interventions [7]. This method was previously applied during the 2009 H1N1 epidemic, caused by an influenza A virus strain [8].

Thus, by retrospectively analyzing the COVID-19 pandemic, it is possible to assess the effectiveness of measures such as vaccination, isolation, and regional variations, identifying those most effective in reducing transmission and severity [9]. The evaluation of risk factors for SARI-related ICU admissions contributes to understanding disease progression, identifying vulnerable groups, and guiding clinical decisions, thereby promoting early interventions [10]. Furthermore, it assists in the allocation of ICU resources by estimating costs during periods of high demand; studies have already contributed to strategies for triage, hospital organization, and intensive therapies [11].

Based on this context, the objective of the present study was to analyze the risk factors for ICU admission at different geographical levels—Brazil, the Southeast region, the state of Minas Gerais, and the municipality of Lavras—as well as to explore hypotheses related to the control measures adopted during the COVID-19 pandemic.

## Materials and Methods

For this analysis, the dataset corresponding to the Severe Acute Respiratory Infection (SARI) Database—including COVID-19 data provided by the OpenDataSUS system [12], which is populated through notification forms submitted to SINAN—was downloaded. Data were extracted on July 9, 2024, covering exclusively the years 2021, 2022, and 2023.

The variables included in the dataset were: municipality, federative unit (FU), sex (male and female), cough, ICU admission, dyspnea, COVID-19 vaccination, influenza vaccination, fever, oxygen saturation <95%, respiratory distress, sore throat, diabetes, and obesity.

Observations with missing information for these variables or classified as “ignored” (code 9) were excluded, reducing the total number of samples from n = 2,567,827 to n = 600,962. After organizing and structuring the database, the sample was filtered by geographical level: Brazil (n = 305,709), the Southeast region (n = 160,418), the state of Minas Gerais (n = 55,350), and the municipality of Lavras (n = 245).

For the modeling process, the class of Generalized Additive Models for Location, Scale, and Shape (GAMLSS), proposed in 2005 by Rigby and Stasinopoulos [13], was applied using the R software [14]. Logistic regression was adopted to model the outcome variable *ICU admission*.

A smoothing function in the form of a cubic spline (cs) was applied to the explanatory variable *age (years)* in order to capture potential nonlinear relationships. Following model specification, a variable selection procedure was performed, excluding variables that were not statistically significant at the 5% level.

## Results

The estimated coefficients for the Severe Acute Respiratory Infection (SARI) ICU admission variable were analyzed separately by geographical region—Brazil, Southeast, Minas Gerais, and Lavras-MG—and are presented in Table 1.

**Table 1.** Estimated coefficients for Intensive Care Unit (ICU) admission due to Severe Acute Respiratory Infection (SARI) in Brazil, the Southeast region, the state of Minas Gerais, and the municipality of Lavras, 2021–2023.

| Variables | Brazil<br>(n=305,709) | Southeast<br>(n=160,418) | Minas Gerais<br>(n=55,350) | Lavras<br>(n=245) | Stat.<br>Sign. |
| --- | --- | --- | --- | --- | --- |
|  | Coef. (S.E.) | Coef. (S.E.) | Coef. (S.E.) | Coef. (S.E.) | p-value |
| **Intercept** | -0.668 (0.021) | -0.689 (0.029) | -0.852 (0.051) | 4.295<br>(1.054) | < 0.05 |
| **Male Sex** | 0.145 (0.007) | 0.136 (0.010) | 0.162 (0.019) | — | < 0.05 |
| **Age (cs)** | 0.001 (0.0001) | -0.001 (0.0002) | 0.002 (0.0005) | — | < 0.05 |
| **Fever** | — | — | — | 0.904<br>(0.3348) | < 0.05 |
| **Cough** | -0.333 (0.008) | -0.258 (0.012) | -0.292 (0.021) | — | < 0.05 |
| **Sore Throat** | -0.127 (0.010) | -0.098 (0.014) | — | 1.418<br>(0.461) | < 0.05 |
| **Dyspnea** | 0.265 (0.010) | 0.236 (0.014) | 0.260 (0.025) | 0.865<br>(0.431) | < 0.05 |
| **Respiratory**<br>**Distress** | 0.249 (0.009) | 0.222 (0.012) | 0.274 (0.021) | — | < 0.05 |
| **Saturation**<br>**<95%** | 0.305 (0.010) | 0.234 (0.013) | 0.189 (0.024) | 1.592<br>(0.398) | < 0.05 |
| Diabetes | 0.110 (0.008) | 0.117 (0.011) | 0.147 (0.020) | — | < 0.05 |
| Obesity | 0.416 (0.011) | 0.381 (0.015) | 0.320 (0.029) | 2.502<br>(0.863) | < 0.05 |
| Covid-19<br>Vaccine | -0.095 (0.008) | -0.112 (0.011) | -0.227 (0.021) | -1.143<br>(0.446) | < 0.05 |
| Influenza<br>Vaccine | -0.257 (0.009) | -0.237 (0.012) | -0.070 (0.022) | -0.669<br>(0.368) | < 0.05 |

Coef.: Estimated Coefficient; S.E.: Standard Error. Cs: Cubic spline, used to model the Age variable.. —: Variable not significant in the model and, therefore, excluded

For Brazil, a 10-fold cross-validation process was performed, which indicated an average accuracy of 67.28% for classifying ICU admissions, with a mean Area Under the Curve (AUC) of 0.6111. The analysis suggests that patients presenting with dyspnea, respiratory distress, and oxygen saturation <95% had a higher risk of ICU admission due to SARI, with respective coefficients of 0.265, 0.249, and 0.305 (*p* < 0.05). Conversely, cough and sore throat were associated with a protective effect, with coefficients of −0.333 and −0.127 (*p* < 0.05). In addition, diabetes and obesity were identified as risk factors, with coefficients of 0.110 and 0.416 (*p* < 0.05), respectively, as was male sex (0.145, *p* < 0.05).

The estimated relationship for age in Brazil is shown in Fig 1, where the risk of SARI-related ICU admission varied according to patient age. The risk increased steadily from 0 to 70 years, followed by a marked decline. This pattern reflects the fact that most of the sample fell within the 40–80 age range, which also corresponded to smaller standard errors.

**Fig 1.**
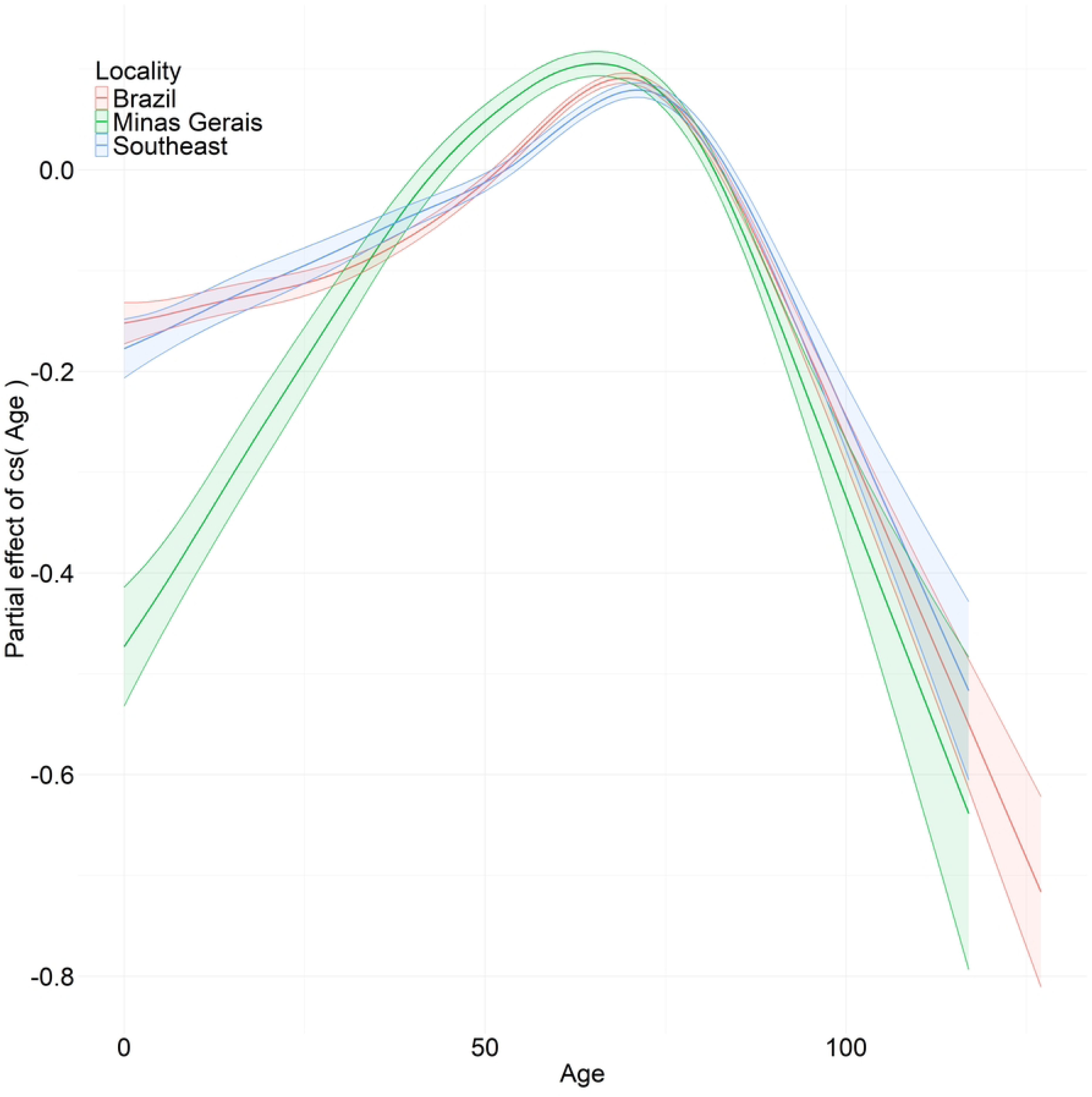
Estimated coefficients for the age variable in modeling the probability of Intensive Care Unit (ICU) admission due to Severe Acute Respiratory Infection (SARI) in Brazil, the Southeast region, and the state of Minas Gerais, 2021–2023.

For Brazil, both COVID-19 and Influenza vaccinations demonstrated a protective effect against SARI-related ICU admission, with coefficients of −0.095 and −0.257 (*p* < 0.05), respectively (Table 1). A statistical analysis was also performed to assess vaccination coverage by age group. As shown in Fig 2A, coverage for both two-dose COVID-19 vaccination and seasonal Influenza vaccination was higher among older patients.

**Fig 2.**
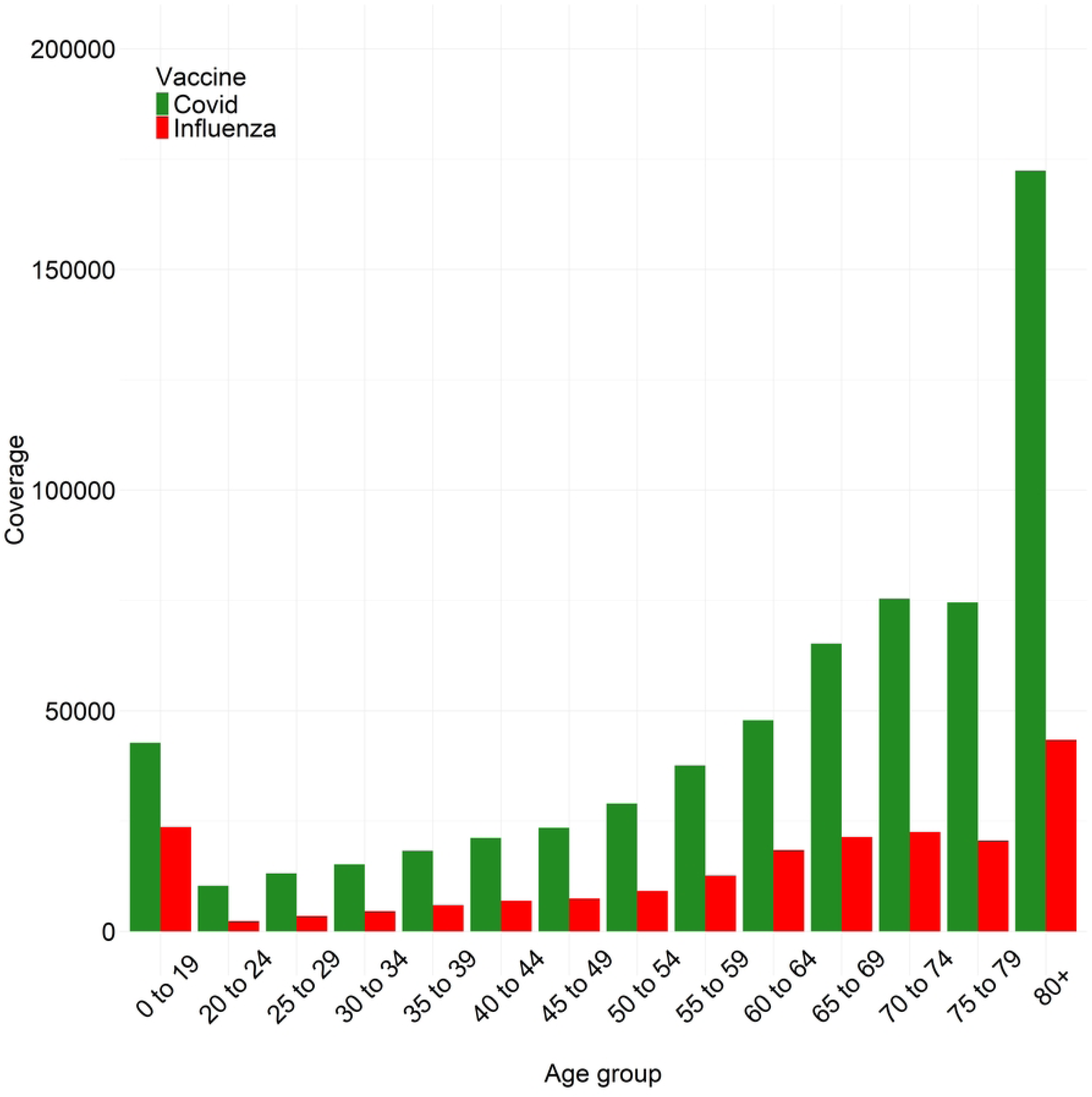
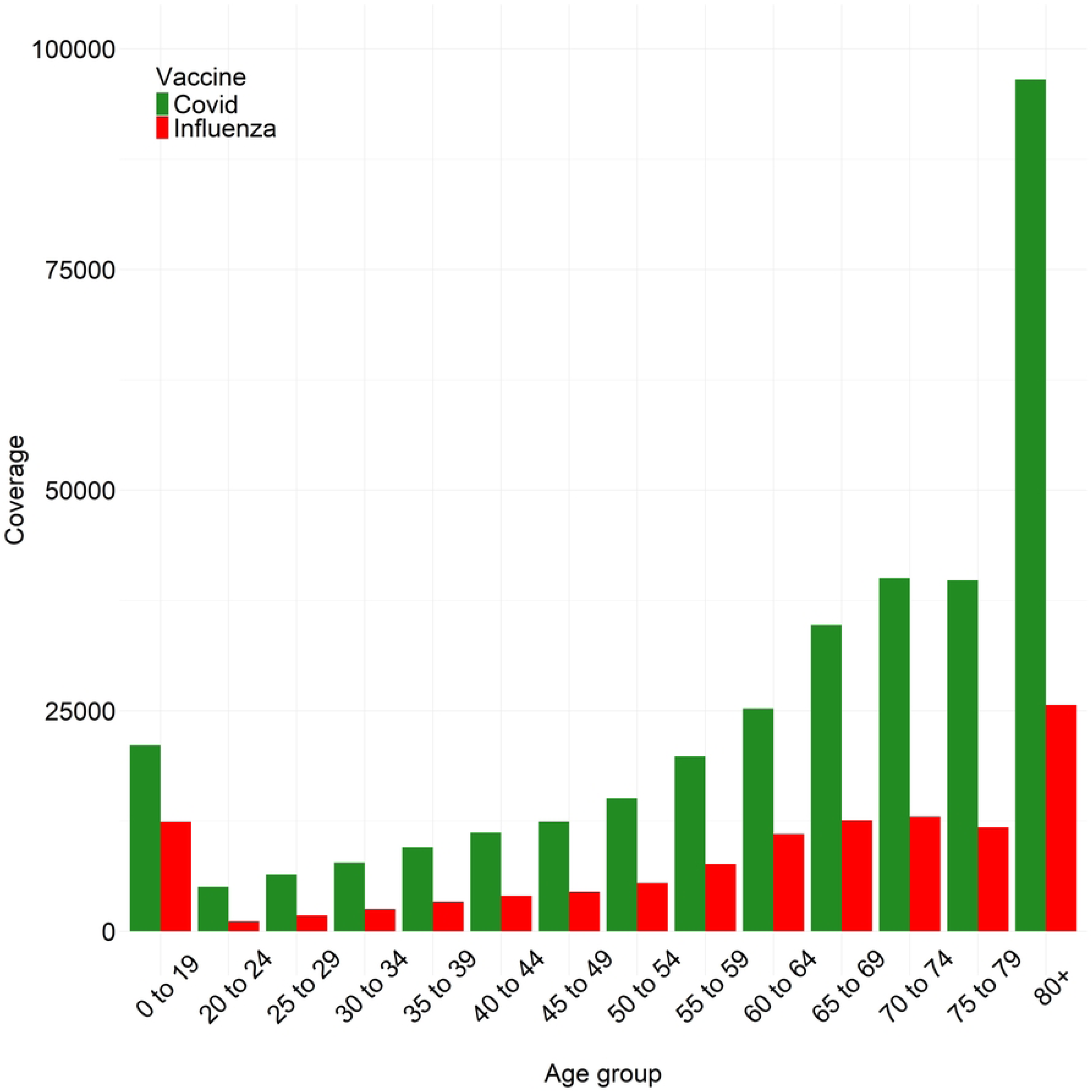
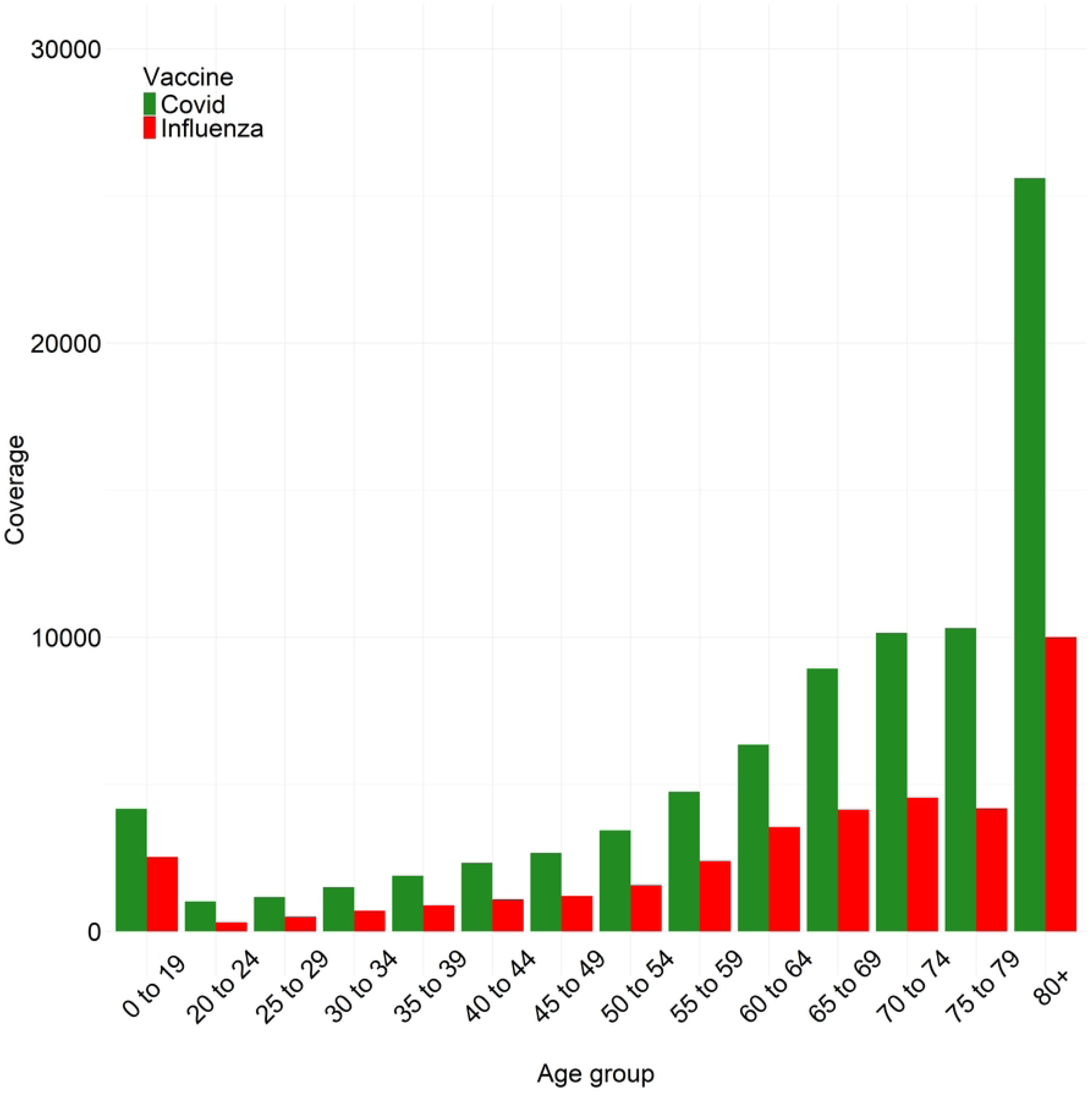
Coverage of two-dose COVID-19 vaccination and seasonal Influenza vaccination by age group in Brazil (A), the Southeast region (B), and the state of Minas Gerais (C), respectively between 2021–2023

For the Southeast region, a 10-fold cross-validation process indicated an average accuracy of 68.00% for classifying ICU admissions, with a mean AUC of 0.5971. The risk relationships observed in the Brazil analysis were maintained in the Southeast: male sex, dyspnea, respiratory distress, oxygen saturation <95%, diabetes, and obesity were all associated with increased risk, with coefficients of 0.136, 0.236, 0.222, 0.234, 0.117, and 0.381 (*p* < 0.05), respectively. Similarly, protective effects were observed for cough, sore throat, COVID-19 vaccination, and Influenza vaccination, with coefficients of −0.258, −0.098, −0.112, and −0.237 (*p* < 0.05), respectively. A graph presented in Fig 1 illustrates the estimated coefficients for SARI-related ICU admission in the Southeast region according to age group from 2021 to 2023.

Given the similar distribution of coefficients by age group in both the Southeast region and Brazil, as shown in Fig 1, an analysis was conducted regarding COVID-19 and Influenza vaccination coverage in the Southeast region by age group (Fig 2B). As in Brazil (Fig 2A), greater COVID-19 vaccination coverage was observed among older patients.

For the state of Minas Gerais, a 10-fold cross-validation process indicated an average accuracy of 72.65% for ICU admission classification, with a mean Area Under the Curve (AUC) of 0.6012. Based on the coefficients presented in Table 1, cough was identified as a protective factor against SARI-related ICU admission in Minas Gerais, with a coefficient of −0.292 (*p* < 0.05). In contrast, dyspnea, respiratory distress, and oxygen saturation <95% were associated with increased risk, with coefficients of 0.260, 0.274, and 0.189 (*p* < 0.05), respectively. Regarding sex and comorbidities, higher risk was observed for males (0.162, *p* < 0.05), as well as for patients with diabetes (0.147, *p* < 0.05) and obesity (0.320, *p* < 0.05). With respect to age in Minas Gerais, the estimated relationship is shown in Fig 1, where the risk of SARI-related ICU admission increased steadily from 0 to 60 years, followed by a decline at older ages, similar to the patterns observed in Brazil and the Southeast region.

COVID-19 and Influenza vaccinations demonstrated a protective effect against ICU admission, with coefficients of −0.227 and −0.070 (*p* < 0.05), respectively (Table 1). An analysis of vaccination coverage by age group was also performed, and the results, presented in Fig 2C, confirmed greater two-dose COVID-19 and Influenza vaccination coverage among older patients.

For the municipality of Lavras (MG), a 10-fold cross-validation process indicated an average accuracy of 73.04% for ICU admission classification, with a mean AUC of 0.8108. According to Table 1, patients presenting with fever, sore throat, dyspnea, and oxygen saturation <95% were more likely to require ICU admission due to SARI, with coefficients of 0.904, 1.418, 0.865, and 1.592 (*p* < 0.05), respectively. Obesity was also identified as a strong risk factor, with a coefficient of 2.502 (*p* < 0.05). For Lavras, the age variable did not reach statistical significance. Both COVID-19 and Influenza vaccinations demonstrated protective effects, with coefficients of −1.143 and −0.669 (*p* < 0.05), respectively.

## Discussion

The present study demonstrated that patients presenting with symptoms such as dyspnea, respiratory distress, and oxygen saturation <95% had an increased risk of ICU admission. These symptoms, considered more severe and systemic, have already been discussed in the literature regarding their association with the severity of SARI manifestations [15]. Certain pathophysiological factors may contribute to greater severity in more symptomatic patients, as the degree of symptomatology may be related both to the patient’s immunological capacity and to the virulence of the etiological agents of SARI [1,16].

Conversely, cough and sore throat were identified as protective factors against SARI-related ICU admission in the analyses for Brazil, the Southeast region, and Minas Gerais. The relationship between these symptoms and SARI severity has not been the focus of most studies, but they may indicate a predominantly upper respiratory tract involvement, suggesting a more localized action of the etiological agents and a lower systemic response from the host [17]. Furthermore, the Brazilian Ministry of Health provides a flowchart for the diagnosis and management of influenza syndrome (IS) and SARI, in which the main distinction between these conditions is based on the presence of severity markers (dyspnea, respiratory distress, oxygen saturation <95%, or exacerbation of pre-existing disease). These markers are characteristic of SARI and frequently lead to hospital or ICU admission. In contrast, signs and symptoms such as low-grade fever, cough, sore throat, myalgia, and headache, in the absence of severity markers, lead to the diagnosis of influenza syndrome, which is managed on an outpatient basis [18].

Despite these findings, and contrary to what was observed for Brazil, the Southeast region, and the state of Minas Gerais, the city of Lavras presented fever and sore throat as risk factors for SARI-related ICU admission. No direct explanation for these results was found in the literature; however, some hypotheses can be proposed. Studies conducted during the pandemic indicated that lethality, epidemiological profiles, and other factors were subject to local influences, including those related to the Human Development Index (HDI), infrastructure, hospital staff, and clinical protocols adopted [19]. In addition, the variability of viral strains may contribute to differences in morbidity and mortality depending on age and geographical groups [16,17].

Considering that Lavras has a Municipal Human Development Index (MHDI) of 0.782 [20]—one of the highest in the state of Minas Gerais—and hosts a federal university medical school [21], it is possible to infer that such structural and institutional factors may have influenced the results obtained. This interpretation is reinforced by comparing the HDI of Lavras (0.782) [20] with that of Minas Gerais (0.774) [22] and Brazil (0.766) [23], supporting the hypothesis that structural differences may underlie the results observed for Lavras. Indeed, municipal-level variables tend to exhibit more homogeneous behavior than those at the state, regional, or national levels.

As expected, a risk association was verified between the presence of comorbidities and the probability of ICU admission, as evidenced by the positive coefficients for diabetes and obesity, consistent with previous studies [16,24,25]. Diabetes is frequently associated with chronic inflammation, which exacerbates the inflammatory response during the pathophysiological process of SARI, in addition to dysfunction of the renin–angiotensin–aldosterone system, potentially aggravating disease progression in diabetic patients [26,27]. Several epidemiological studies have also identified diabetes as an independent risk factor for unfavorable prognosis in SARI patients, with significantly higher mortality and complication rates compared to non-diabetic patients [28]. Similarly, obesity—often associated with other cardiovascular risk factors such as diabetes and hypertension—has been linked to more severe SARI outcomes. This may be partially explained by the immunological and inflammatory dysfunction associated with excess adipose tissue [29,30]. Despite its frequent association with other cardiovascular risk conditions, obesity was identified as an independent risk factor for ICU admission and increased SARI severity [31].

Male sex was also associated with an increased risk of ICU admission. This can be explained by a more efficient innate immune response in females, possibly related to the effects of female hormones (estrogens and progesterone), which exert protective effects against viral infections. In contrast, male hormones (testosterone) increase the expression of the Angiotensin-Converting Enzyme 2 (ACE2) receptor, which, particularly in the context of COVID-19, facilitates the entry of SARS-CoV-2 into cells, increasing viral load in men and delaying viral clearance [32]. Moreover, men tend to present a higher prevalence of comorbidities such as obesity, cardiovascular diseases, and hypertension, which are established risk factors for greater SARI severity [33]. Social factors may also contribute, as risk behaviors such as smoking and alcohol consumption are more common among men, along with lower adherence to public health measures such as mask use and hand hygiene—factors shown to increase exposure and infection severity [34].

The graphs of the estimated age relationships in Fig 1 show higher standard errors at the extremes, which may be explained by the smaller number of very elderly individuals in the sample. Nevertheless, this does not preclude the unexpected finding of a reduction in ICU admission risk among older patients, particularly from the age of 60 onward. Previous studies have demonstrated a strong association between advanced age and increased SARI severity, especially in the epidemiological context of the COVID-19 pandemic. This is linked to age-related immune dysfunction, characterized by an ineffective and dysregulated immune response, with a reduction in the population of naïve T and B cells and an increase in monocytes and senescent T cells, resulting in a prolonged and potentially pathological inflammatory response [35]. In addition, elderly patients may exhibit increased ACE2 receptor expression, facilitating viral replication, as discussed earlier, combined with comorbidities associated with aging, such as cardiovascular disease and diabetes, which further contribute to disease severity [36].

However, a protective association was identified between COVID-19 and Influenza vaccination and the probability of SARI-related ICU admission. A higher prevalence of vaccination—particularly two-dose COVID-19 vaccination and seasonal Influenza vaccination—was observed among patients aged >55 years, suggesting that this may explain the results obtained for the age variable. This finding is consistent with other studies, which have highlighted the protective effect of COVID-19 vaccination against increased SARI severity [15], along with the fact that elderly patients were prioritized for COVID-19 vaccination [37].

## Conclusion

This study concluded that patients presenting with systemic symptoms such as dyspnea, oxygen saturation <95%, and respiratory distress had a higher probability of SARI-related ICU admission. However, local factors—such as the Municipal Human Development Index (MHDI), ICU bed availability, and other structural aspects—may influence the risk that less severe upper airway symptoms could lead to the same outcome. The presence of diabetes, obesity, and male sex was also associated with an increased likelihood of ICU admission, consistent with findings reported in the literature. Unexpectedly, age ≥60 years was associated with a lower risk of ICU admission, a result that may be explained by higher COVID-19 vaccination coverage among older individuals during the analyzed period.

## Data Availability

The data are available and may be accessed by any interested party upon request.

## Acknowledgments

This research was made possible through access to data provided by the OpenDataSUS/SINAN system, referring to cases of Severe Acute Respiratory Infection (SARI) in Brazil. The author acknowledges funding from the Institutional Program for Scientific Initiation Scholarships (PIBIC) of the Fundação de Amparo à Pesquisa do Estado de Minas Gerais (FAPEMIG), through the scholarship granted to support the development of this work at the Universidade Federal de Lavras (UFLA). The support of the Coordenação de Aperfeiçoamento de Pessoal de Nível Superior (CAPES) and the Conselho Nacional de Desenvolvimento Científico e Tecnológico (CNPq), whose investments in science and researcher training were fundamental to the accomplishment of this study, is also gratefully recognized. Department of Foundational Sciences, School of Dental Medicine, East Carolina University (ECU), Greenville, North Carolina, USA.

## References

1. Araujo KLR de, Aquino ÉC de, Silva LLS da, Ternes YMF. Fatores associados à Síndrome Respiratória Aguda Grave em uma Região Central do Brasil. Ciência & Saúde Coletiva [Internet]. 2020 Oct [cited 2025 Sep 28];25(suppl 2):4121–30. Available from: https://www.scielo.br/j/csc/a/vyW3LvH4KB38LQq4qvGVpPs/

2. Ministério da Saúde. Protocolo De Tratamento De Influenza: 2017 [Internet]. Brasília: Ministério da Saúde. 2018 [cited 2025 Sep 28]. Available from: https://bvsms.saude.gov.br/bvs/publicacoes/protocolo_tratamento_influenza_2017.pdf

3. Silva AP de SC, Maia LT de S, Souza WV de. Síndrome Respiratória Aguda Grave Em Pernambuco: Comparativo Dos Padrões Antes E Durante a Pandemia De COVID-19. Ciência & Saúde Coletiva [Internet]. 2020 Sep 30 [cited 2021 May 14];25(suppl 2):4141–50. Available from: https://scielosp.org/article/csc/2020.v25suppl2/4141-4150/pt/

4. Ministério da Saúde. Coronavírus Brasil [Internet]. covid.saude.gov.br. 2024. Available from: https://covid.saude.gov.br/

5. Paiva KM de, Hillesheim D, Rech CR, Delevatti RS, Brown RVS, Gonzáles AI, et al. Prevalência E Fatores Associados À SRAG Por COVID-19 Em Adultos E Idosos Com Doença Cardiovascular Crônica. Arquivos Brasileiros De Cardiologia [Internet]. 2021 Jun 10;117(5). Available from: https://www.scielo.br/j/abc/a/bM5Z6WDY83RbQyfLbkQZ4vC/?lang=pt

6. Americo Xavier L, Paes Santos Neto CA, Pinheiro Rodrigues R, da Silva Sousa Júnior A, Mourão de Sousa A. Vista Do Síndrome Respiratória Aguda Grave Por Covid-19: Internações Em Unidade De Terapia Intensiva Nas Regiões De Saúde Do Estado Do Pará, 2020-2022 [Internet]. Redeunida.org.br. 2020 [cited 2025 Sep 28]. Available from: https://revista.redeunida.org.br/index.php/rede-unida/article/view/4285/1375

7. James J Schlesselman. Case Control studies: design, conduct, analysis. Vol. 2. New York, Oxford University Press; 1982.

8. Pangratz Bedretchuk G, Hubie APS, Cavalli LO. Vista Do Perfil Sociodemográfico Do Paciente Acometido Por Síndrome Respiratória Aguda grave: Um Estudo Retrospectivo De Nove Anos. FAG Journal of Health [Internet]. 2019 Dec 20 [cited 2022 Oct 16];1(4):67–78. Available from: https://pdfs.semanticscholar.org/a3e5/757bcd40223d88f869bf99f7938e9bc8aea4.pdf

9. Dean NE, Hogan JW, Schnitzer ME. Covid-19 Vaccine Effectiveness and the Test-Negative Design. New England Journal of Medicine [Internet]. 2021 Sep 8;385(15). Available from: https://www.nejm.org/doi/full/10.1056/NEJMe2113151

10. Grasselli G, Greco M, Zanella A, Albano G, Antonelli M, Bellani G, et al. Risk Factors Associated with Mortality among Patients with COVID-19 in Intensive Care Units in Lombardy, Italy. JAMA Internal Medicine [Internet]. 2020 Oct 1 [cited 2025 Sep 28];180(10):1345. Available from: https://jamanetwork.com/journals/jamainternalmedicine/fullarticle/2768601

11. Cummings MJ, Baldwin MR, Abrams D, Jacobson SD, Meyer BJ, Balough EM, et al. Epidemiology, Clinical course, and Outcomes of Critically Ill Adults with COVID-19 in New York City: a Prospective Cohort Study. The Lancet [Internet]. 2020 Jun [cited 2025 Sep 28];395(10239):1763–70. Available from: https://www.thelancet.com/journals/lancet/article/PIIS0140-6736(20)31189-2/fulltext

12. Ministério da Saúde. SRAG 2019 a 2025 - Banco De Dados De Síndrome Respiratória Aguda Grave - Incluindo Dados Da COVID-19 - OPENDATASUS [Internet]. Opendatasus. 2025. Available from: https://opendatasus.saude.gov.br/dataset/srag-2021-a-2024

13. Rigby RA, Stasinopoulos DM. Generalized Additive Models for location, Scale and Shape (with discussion). Journal of the Royal Statistical Society: Series C (Applied Statistics) [Internet]. 2005 Jun;54(3):507–54. Available from: https://rss.onlinelibrary.wiley.com/doi/abs/10.1111/j.1467-9876.2005.00510.x

14. R Core Team. R: a Language and Environment for Statistical Computing [Internet]. R Foundation for Statistical Computing. 2025 [cited 2025 Sep 28]. Available from: https://www.r-project.org/

15. Batista Silva A, Fernandes Campos Coelho H, Roseane Queiroz de Paiva Faria A, Rocha Carneiro R, Bezerra Luna Lima CM. Fatores Associados Ao Óbito Em Pacientes Internados Por Síndrome Respiratória Aguda Grave Na Paraíba. Journal of Nursing and Health [Internet]. 2024 Jul 16 [cited 2025 Sep 28];14(1):e1424474. Available from: https://periodicos.ufpel.edu.br/index.php/enfermagem/article/view/24474

16. Kfuri T, Mafra A, Stobbe J, Dos R, Rabello S, Lindemann I, et al. A Síndrome Respiratória Aguda Grave Na Pessoa Idosa No Contexto Da Pandemia Da covid-19 E Seus Fatores Associados. Revista Brasileira De Geriatria E Gerontologia [Internet]. 2023 [cited 2025 Sep 28];26(1). Available from: https://www.rbgg.com.br/edicoes/v26/RBGG%20v26%20PORT_2022-0158_PRONTO.pdf

17. Marques Vieira Izecksohn M, da Costa Leite I, Paiva Daumas R, Esteves Siqueira Rodrigues R. Fatores Associados À Ocorrência De Síndrome Respiratória Aguda Grave (SRAG) Entre Casos De COVID-19. Ciência & Saúde Coletiva [Internet]. 2025 Jan 1 [cited 2025 Sep 28];30(7). Available from: https://www.scielo.br/j/csc/a/rq5f5dpknnsyP8PWrcpqwwh/?format=html&lang=pt

18. Ministério da Saúde. Síndrome Gripal/SRAG:Classificação De Risco E Manejo Do Paciente [Internet]. Biblioteca Virtual De Saúde; 2013 [cited 2025 Sep 28]. Available from: https://bvsms.saude.gov.br/bvs/folder/sindrome_gripal_sindrome_respiratoria_aguda_grave.pdf

19. Goulart LFM, Ricardo de Oliveira L, Domingos dos Santos Cintra Lima L, Estevam Simonato L, Fernanda Rodrigues Frias D. Perfil Epidemiológico Dos Casos De Síndrome Respiratória Aguda Grave No Estado De Minas Gerais, Brasil, 2020 a 2021. Vigilância Sanitária Em Debate [Internet]. 2023 Aug 3 [cited 2025 Sep 28];11(e02062):1. Available from: https://visaemdebate.incqs.fiocruz.br/index.php/visaemdebate/article/view/2062

20. Brasil. Lavras (MG) | Cidades E Estados | IBGE [Internet]. www.ibge.gov.br. 2010 [cited 2025 Sep 28]. Available from: https://www.ibge.gov.br/cidades-e-estados/mg/lavras.html

21. Reis F. Medicina - Faculdade De Ciências Da Saúde (FCS) [Internet]. Universidade Federal De Lavras. 2015 [cited 2025 Sep 28]. Available from: https://fcs.ufla.br/graduacao/medicina-bacharelado

22. Brasil. Minas Gerais | Cidades E Estados | IBGE [Internet]. www.ibge.gov.br. 2021 [cited 2025 Oct 9]. Available from: https://www.ibge.gov.br/cidades-e-estados/mg.html

23. UDH. Painel IDHM | United Nations Development Programme [Internet]. UNDP. 2021 [cited 2025 Oct 9]. Available from: https://www.undp.org/pt/brazil/desenvolvimento-humano/painel-idhm

24. Pereira Corrêa V, Reis da Silveira E, Rodrigues da Luz J, Vanti da Rocha B. Fatores Relacionados Ao Óbito Por COVID-19: Estudo Transversal Em Município Catarinense. Revista de Saúde Coletiva da UEFS [Internet]. 2024 Mar 31 [cited 2025 Sep 28];14(1):e9382. Available from: https://ojs3.uefs.br/index.php/saudecoletiva/article/view/9382

25. Alves DAN da S, Nascimento GILA, Castanha ER, Luna JEL, Sobral EFM, Brandão WA, et al. Prevalência De Comorbidades Na Síndrome Respiratória Aguda Grave Em Pacientes Acometidos Por COVID-19 E Outros Agentes Infecciosos. Research, Society and Development [Internet]. 2020 Dec 1 [cited 2025 Sep 28];9(11):e70791110286. Available from: https://rsdjournal.org/index.php/rsd/article/view/10286

26. Bonyek-Silva I, Machado AFA, Cerqueira-Silva T, Nunes S, Silva Cruz MR, Silva J, et al. LTB4-Driven Inflammation and Increased Expression of ALOX5/ACE2 during Severe COVID-19 in Individuals with Diabetes. Diabetes [Internet]. 2021 Sep;70(9):2120–30. Available from: https://pubmed.ncbi.nlm.nih.gov/34417262/

27. Obukhov AG, Stevens BR, Prasad R, Li Calzi S, Boulton ME, Raizada MK, et al. SARS-CoV-2 Infections and ACE2: Clinical Outcomes Linked with Increased Morbidity and Mortality in Individuals with Diabetes. Diabetes [Internet]. 2020 Jul 15 [cited 2025 Sep 28];69(9):1875–86. Available from: https://diabetesjournals.org/diabetes/article/69/9/1875/39428/SARS-CoV-2-Infections-and-ACE2-Clinical-Outcomes

28. Shang J, Wang Q, Zhang H, Wang X, Wan J, Yan Y, et al. The Relationship between Diabetes Mellitus and COVID-19 Prognosis: a Retrospective Cohort Study in Wuhan, China. The American Journal of Medicine [Internet]. 2020 Jul [cited 2020 Nov 9];134(1). Available from: https://www.sciencedirect.com/science/article/pii/S0002934320305325

29. Rebello CJ, Kirwan JP, Greenway FL. Obesity, the Most Common Comorbidity in SARS-CoV-2: Is Leptin the link? International Journal of Obesity [Internet]. 2020 Jul 9 [cited 2025 Sep 28];44(9):1810–7. Available from: https://www.nature.com/articles/s41366-020-0640-5

30. Denson JL, Gillet AS, Zu Y, Brown M, Pham T, Yoshida Y, et al. Metabolic Syndrome and Acute Respiratory Distress Syndrome in Hospitalized Patients with COVID-19. JAMA Network Open [Internet]. 2021 Dec 22 [cited 2022 Jan 25];4(12):e2140568. Available from: https://jamanetwork.com/journals/jamanetworkopen/fullarticle/2787394

31. Kim L, Garg S, O’Halloran A, Whitaker M, Pham H, Anderson EJ, et al. Risk Factors for Intensive Care Unit Admission and In-hospital Mortality among Hospitalized Adults Identified through the U.S. Coronavirus Disease 2019 (COVID-19)-Associated Hospitalization Surveillance Network (COVID-NET). Clinical Infectious Diseases [Internet]. 2020 Jul 16 [cited 2020 Aug 7];72(9). Available from: https://academic.oup.com/cid/article/72/9/e206/5872581

32. Viveiros A, Rasmuson J, Vu J, Mulvagh SL, Yip CYY, Norris CM, et al. Sex Differences in COVID-19: Candidate pathways, Genetics of ACE2, and Sex Hormones. American Journal of Physiology-Heart and Circulatory Physiology. 2021 Jan 1;320(1):H296–304.

33. Iaccarino G, Grassi G, Borghi C, Carugo S, Fallo F, Ferri C, et al. Gender Differences in Predictors of Intensive Care Units Admission among COVID-19 patients: the Results of the SARS-RAS Study of the Italian Society of Hypertension. Shimosawa T, editor. PLOS ONE [Internet]. 2020 Oct 6;15(10):e0237297. Available from: https://pubmed.ncbi.nlm.nih.gov/33022004/

34. MacArthur TA, Goswami J, Ramachandran D, Price-Troska TL, Lundell KA, Ballinger BA, et al. Estradiol and Dihydrotestosterone Levels in COVID-19 Patients. Mayo Clinic Proceedings [Internet]. 2023 Jan 17 [cited 2025 Sep 28];98(4):559–68. Available from: https://pmc.ncbi.nlm.nih.gov/articles/PMC9842620/

35. Pedroso RB, Torres L, Ventura LA, Camatta GC, Mota C, Mendes AC, et al. Rapid Progression of CD8 and CD4 T Cells to Cellular Exhaustion and Senescence during SARS-CoV2 Infection. Journal of Leukocyte Biology [Internet]. 2024;116(6):1385–97. Available from: https://pubmed.ncbi.nlm.nih.gov/39298288/

36. Farshbafnadi M, Kamali Zonouzi S, Sabahi M, Dolatshahi M, Aarabi MH. Aging & COVID-19 susceptibility, Disease severity, and Clinical outcomes: the Role of Entangled Risk Factors. Experimental Gerontology [Internet]. 2021 Oct [cited 2025 Sep 28];154:111507. Available from: https://pubmed.ncbi.nlm.nih.gov/34352287/

37. Pagno M. Entenda a Ordem De Vacinação Contra a Covid-19 Entre Os Grupos Prioritários [Internet]. Ministério Da Saúde. 2022 [cited 2025 Oct 9]. Available from: https://www.gov.br/saude/pt-br/assuntos/noticias/2021/janeiro/entenda-a-ordem-de-vacinacao-contra-a-covid-19-entre-os-grupos-prioritarios

